# Addressing tuberculosis in artisanal and small-scale mining activities in sub-Saharan Africa: meta-analysis and a call for actions

**DOI:** 10.1101/2023.08.24.23294543

**Authors:** Daniel Garhalangwanamuntu Mayeri, Richard Mbusa Kambale, Patrick Musole Bugeme, Gaylor Amani Ngaboyeka, Charles Mushagalusa, Franck Mugisho Zahinda, Jacques L. Tamuzi, Patrick DMC Katoto

## Abstract

**Background:** Tuberculosis (TB) is a significant health issue in sub-Saharan African (SSA) countries, and artisanal mining (AM) may be a contributing factor. However, no systematic review has investigated the association between AM and TB in SSA. Therefore, this study aims to assess the burden of TB among artisanal miners in SSA.

**Methods:** We conducted a comprehensive search of PubMed, Medline-OVID, EMBASE, and Scopus databases for studies on AM and TB published up to January 25, 2022. We presented the findings of seven studies that met our inclusion criteria narratively and through figure synthesis, and used inverse-variance weighted random-effects models to combine effect estimates for meta-analysis.

**Results:** The overall prevalence of TB among artisanal workers was estimated to be 15% (95%CI: 8, 23), with higher rates in high TB burden countries (19%, 95%CI: 11, 28) compared to upper-moderate burden countries (8%, 95%CI: 3, 19. Further, exposure to silica dust, a common byproduct of AM, significantly increased the incidence of TB, with a pooled relative risk of 2.19 (95% CI: 1.77, 2.71). Additionally, we found that a higher number of artisanal miners in Ghana was associated with a reduction in TB incidence.

**Conclusion:** Our findings suggest that exposure to silica dust in AM is a neglected but a significant risk factor for TB in SSA. More studies and efforts are needed to address this threat to TB control.

## Introduction

Millions of deaths related to air pollution in artisanal mines and pulmonary tuberculosis (PTB) occur annually in low-income countries [1,2]. This underreported epidemic contributes heavily to the global burden of disease each year and is one of the top five causes of death in most Sub-Saharan countries (SSA). According to the World Health Organization (WHO) Global Health Observatory, living or working in an unhealthy environment was associated with 13.7 million deaths in 2016 representing 24.3% of all yearly deaths [3]. The Lancet Commission on Pollution and Health 2017 stressed the fact that people from low- and middle-income countries (LMICs), disproportionately experience the burden of adverse effects due to air pollution [3].

The prevalence of tuberculosis (TB) has not been effectively controlled in most countries due to challenges posed by civil war, weak health systems, and national TB programs focusing only on diagnosis and treatment [11,12]. Several countries in Africa and beyond frequently report cases of TB, particularly those with active small and large-scale mining operations, such as South Africa, India, and China. These countries are recognized as having a high burden of TB. [6][7, 8][9]. In addition, in the past decade, the Democratic Republic of Congo (DRC), a country with a high burden of TB, HIV, and ASM, has seen an increase in TB-related deaths among HIV-negative individuals, from 57% to 61%[10]. This shift in TB control in DRC is unexpected and indicates the need to investigate other potential risk factors, including environmental factors.

Based on our understanding of the impact of tobacco smoking, diesel exhaust particles, and fine particulate matter with a diameter less than 2.5 μm, there is substantial evidence indicating a significant association between exposure to these pollutants and adverse health effects, particularly the development of tuberculosis (TB). Pollutants and *Mycobacterium tuberculosis* have been found to generate oxidative stress and heightened systemic inflammation, exacerbating their negative health impacts[4]. Similarly, exposure to silica dust has been associated with an elevated risk of TB. Silica, which accounts for more than 90% of the Earth’s crust (SiO2), is the second most common silicate mineral. Individuals residing in mining zones, as well as those employed in construction, pottery, and other industries, may have increased environmental or occupational exposure to silica or silicates. However, the risk of developing active TB is more than three times higher in individuals with silicosis (known as silico-TB), with HIV-positive silicosis patients having a higher incidence of TB compared to other HIV-positive patients [5].

Unfortunately, outside of industrial mining exploitation, there is limited knowledge about the relationship between exposure to pollution from the growing ASM in SSA and its impact on TB control, primarily due to a lack of personal ground-level exposure measurements and regulations in this industry. Therefore, the purpose of this review is to systematically summarize the literature on the burden of tuberculosis in the SSA artisanal mining setting to strengthen ASM sector regulations and design protective interventions to reduce the burden of TB among ASM workers and their communities.

## Materials and Methods

### Search strategy and selection criteria

We searched PubMed and Medline-OVID, EMBASE, and Scopus databases using a combination of terms and derived keywords, and including variations of the following terms: “artisanal”, “artisanal mining”, “health”, “and health effects”, “tuberculosis”, “prevalence”, “risk factor”(See Table 6 for full search strategy). In the MEDLINE search, the names of each Sub-Saharan African country (“United Nations - Sub-Saharan Africa,” n.d.) as well as regions (e.g., East-Africa) were included to increase search sensitivity. We also searched for eligible studies in the WHO database, national agencies, African Index Medicus, abstracts at scientific conferences (e.g., Pan African Thoracic Society), and other relevant grey literature. No language restriction was applied; the timeframe of the search included all records from the electronic database inception to January 24, 2022.

We included all the studies from one of the SSA and that evaluated TB in the context of artisanal mining. Thus, we used the PECO framework for inclusion criteria [14] as follows:

– **Population**: Adults and the elderly who were exposed to dust in artisanal mining and developed TB.
– **Exposure**: studies of individuals with respiratory exposure to dust in artisanal mines inferred from occupation.
– **Comparators**: studies reporting comparative effect estimates, specifically case-control or cohort studies reporting risk, rate, or odds across groups exposed to dust in mines (including binary comparisons of exposed/unexposed).
– **Outcome:** studies reporting incident active pulmonary TB, with or without extra pulmonary tuberculosis. TB diagnosis was made on histological or microbiological grounds, or an explicit combination of clinical assessment, radiology and/or response to treatment. Furthermore, TB was diagnosed after mining activities.

Two reviewers (DGM and PBM) evaluated the eligibility of studies. In cases of discrepancy, a third reviewer (PK) provided resolution of disagreements. For exposure, we extracted the following information: reference, country/city/town, period, context, design (analytic method), type of mining and other relevant considerations (comparison with international standards, etc.). Data included first author, publication year, study country, study date, study design (effect measure), study population and data sources for comparison group, sample size, TB diagnosis methods, Exposure assessment and summary of main conclusions [estimates of associations (95% confidence intervals).

### Risk of bias assessment

For quality and risk of bias assessment, we used the Newcastle-Ottawa Scale (NOS) (“Ottawa Hospital Research Institute,” n.d.), as recommended by the Cochrane Non-Randomized Studies Methods Working Group (“Cochrane Handbook for Systematic Reviews of Interventions,” n.d.). The NOS uses an eight-item rating system to evaluate the method of selecting participants, exposure/outcome assessment, and comparability among study groups. It has specific formats for cohort and case-control studies. We used the modified form for cross-sectional studies and case-control studies.

Two reviewers (DGM and PBM) independently performed the quality assessment and a third reviewer (PK) resolved disagreements.

### Data analysis

We performed a meta-analysis using RevMan 5.4 and Stata 16 software. If studies were homogenous, we used a random-effects model to synthesize all data regardless of heterogeneity between the pooled studies. Adjusted analysis using the effect estimate reported in the study was reported. If only crude data were available, the rate ratio and 95% CIs dichotomous data were computed and pooled with the adjusted estimates. Statistical heterogeneity was evaluated with the Chi2 test with significance set at P value < 0.1. We measured the degree of heterogeneity among the studies by the I2 statistic. Subgroup analyses were performed to assess the influence of study period and study design on the main outcome. Furthermore, we built a meta-regression model including the number of ASM and TB burden in countries where studies were conducted. Egger’s and Begg’s rank correlation tests were used to explore publication bias. Sensitivity analysis was not performed due to the small number of studies in each subgroup.

ArcGIS software was used to generate maps of SSA, which were colored on one side for countries with a high burden of TB and high levels of artisanal mining activities, and on the other side for countries with a high TB burden and artisanal mining activities. Studies investigating the association between artisanal mining and TB were also reported. [15,16]. This systematic review was reported according to the Preferred Reporting Items for Systematic Reviews and Meta-analyses (PRISMA) guideline [17,18].

## 3. Results

### 3.1 Study selection

The electronic research identified a total of 1,597 papers, of which 442 were screened according to the inclusion criteria. Four hundred and seventeen of them were excluded and only 25 full texts were assessed for eligibility. Finally, 7 were eligible for data extraction and analysis and consequently included in this review (Figure 1).

**Fig. 1.**
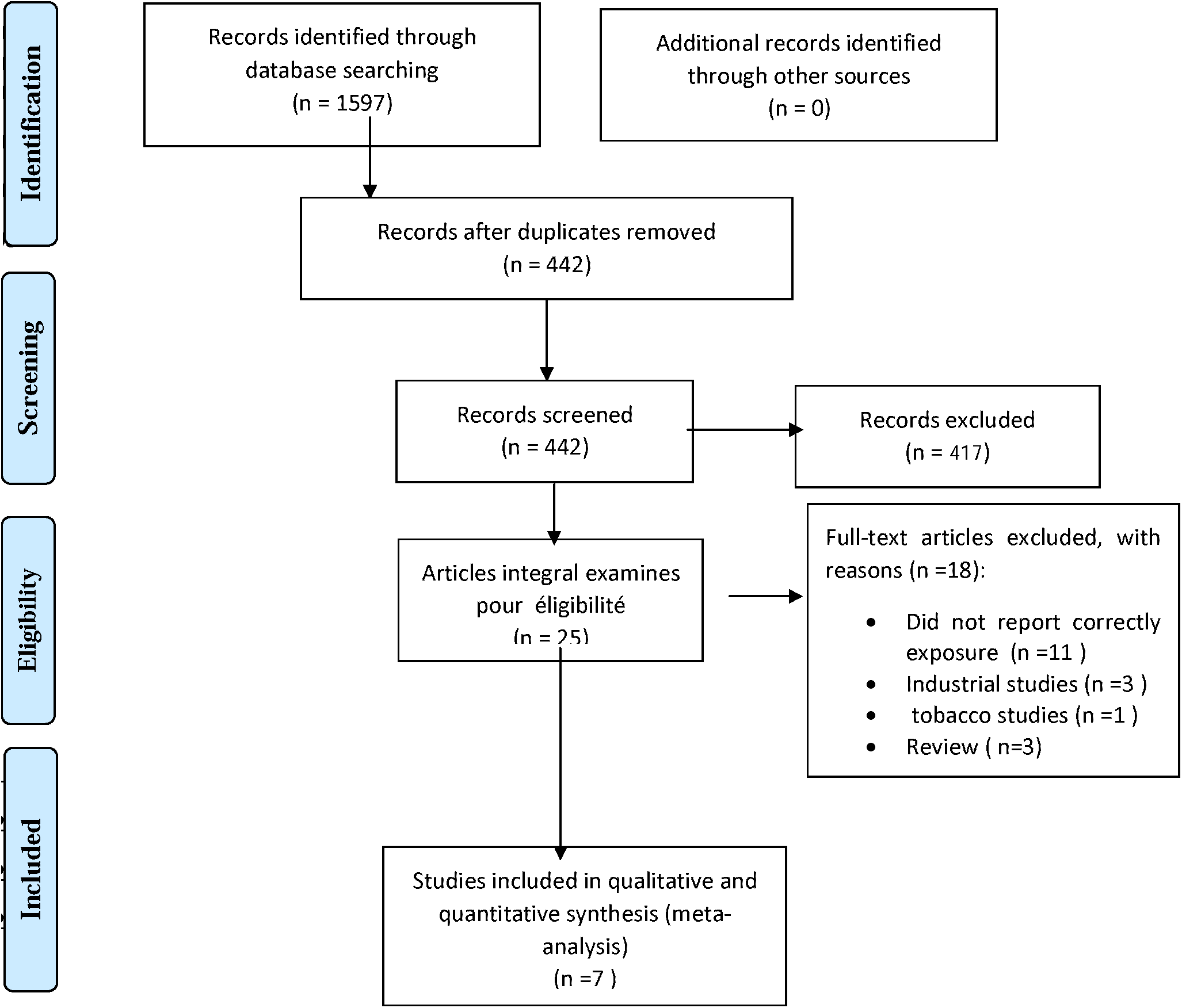
PRISMA flow diagram for selection of studies. SSA: Sub-Saharan Africa.

### 3.2 Overall Study characteristics

Among the seven studies, two were cohort studies [19,20], one was a case-control study [5] and four others were cross-sectional [21–24]. (Table 1 and Figure 2). Only 3 countries (South Africa, Ghana and Malawi) out of the 46 that constitutes SSA region provided data on TB and artisanal mining activities. One from the Western part of Africa, and two from the Southern Africa. South Africa for itself is represented by more 57% of the available studies [5, 19–21]. All the 3 countries (South Africa, Ghana, and Malawi) where the studies have been conducted have a high burden of TB in SSA. The publication year ranged from 1994 to 2021. All the cases were either working in artisanal mines or have had a history of artisanal mining. One of the studies [24] reported rifampicin resistance associated to mining activities.

**Table 1:**
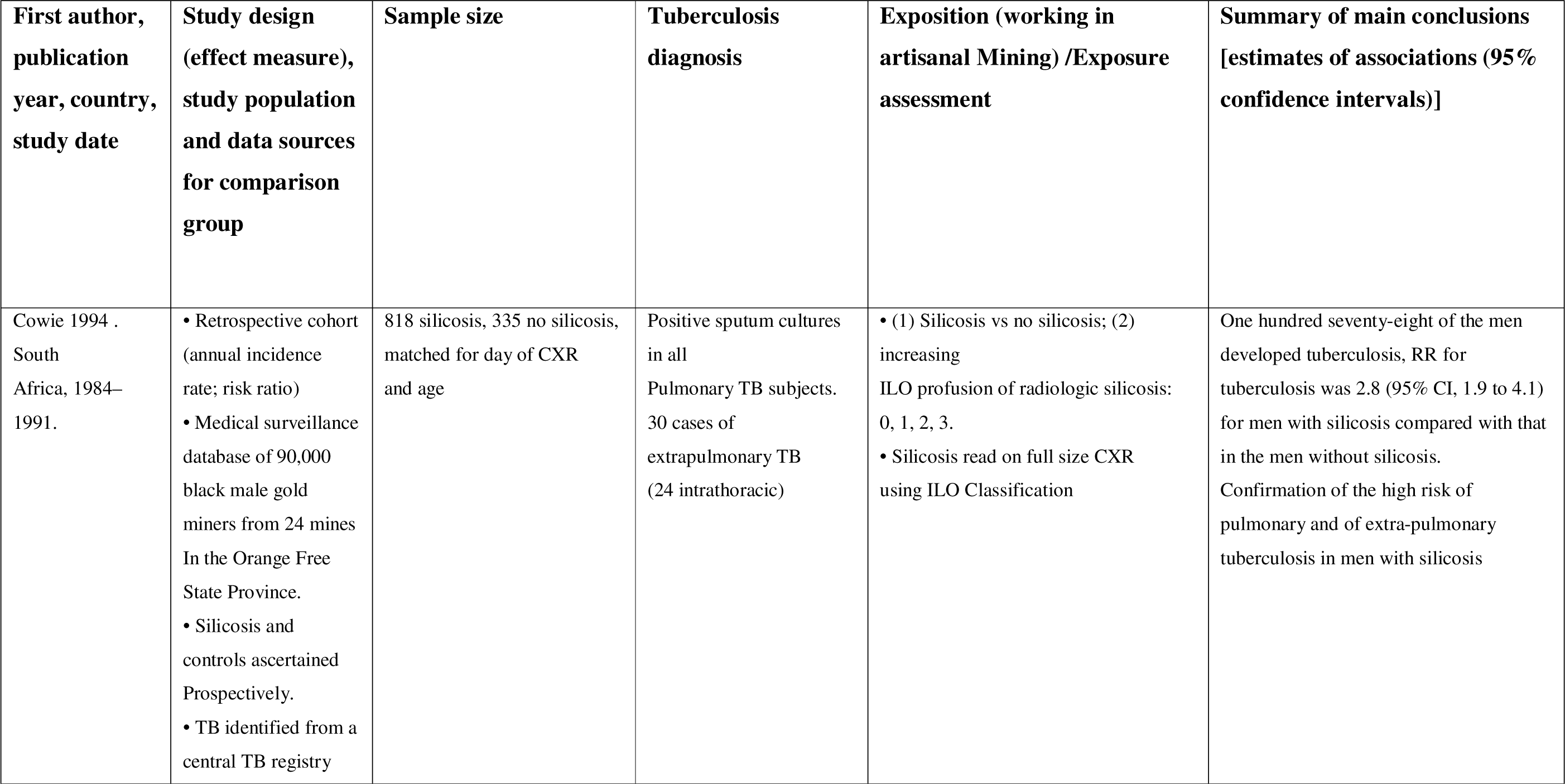
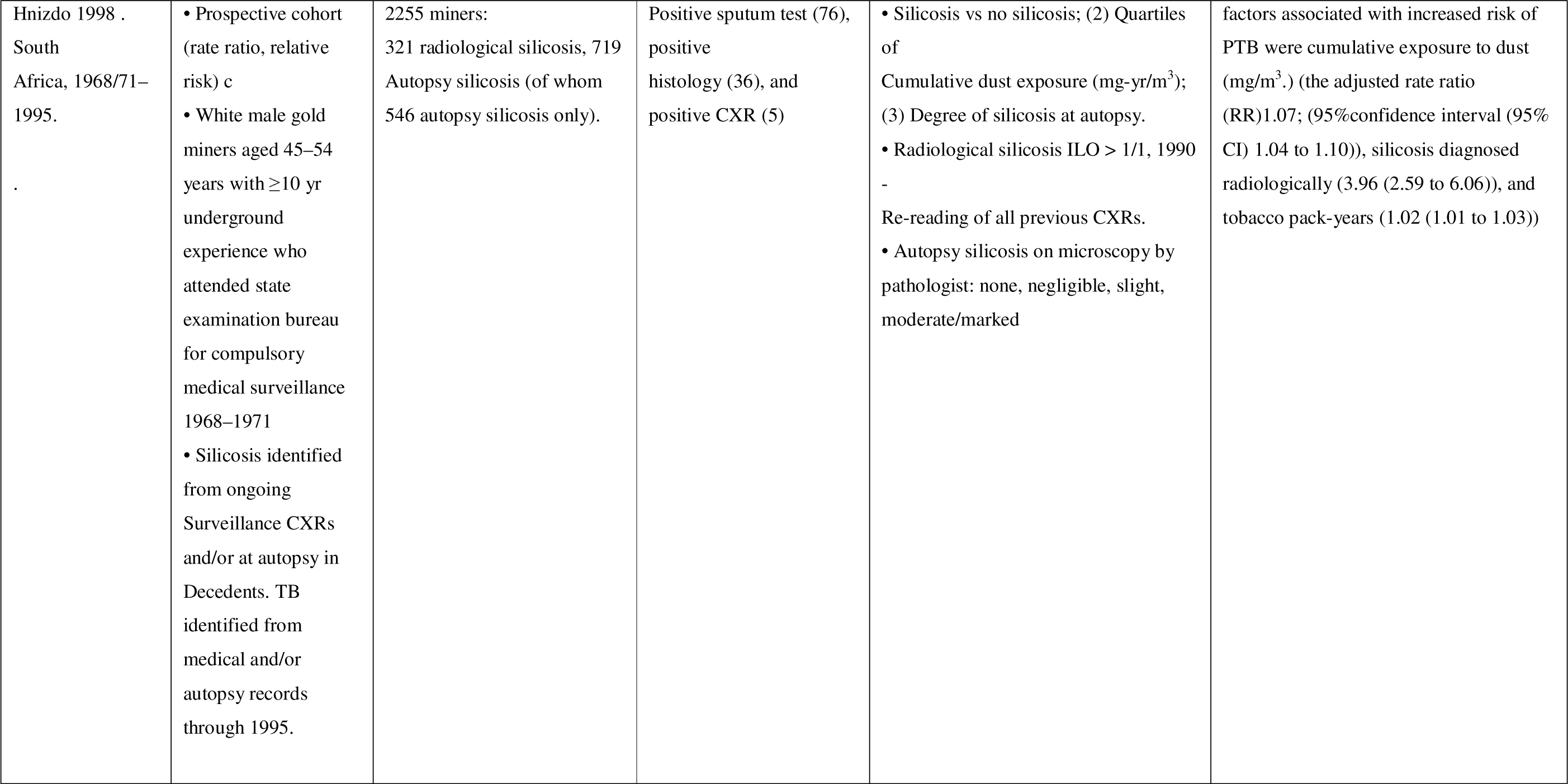
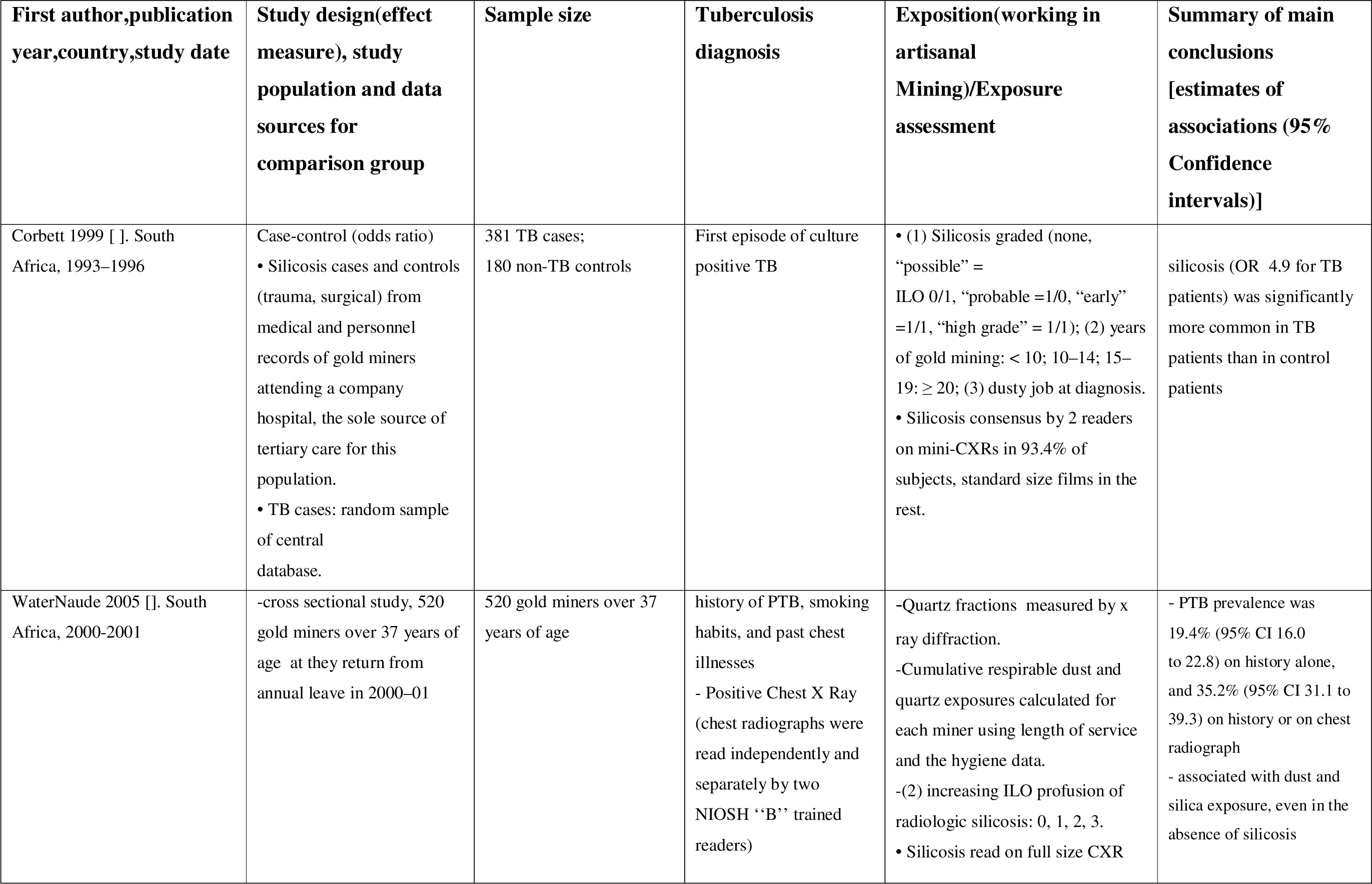
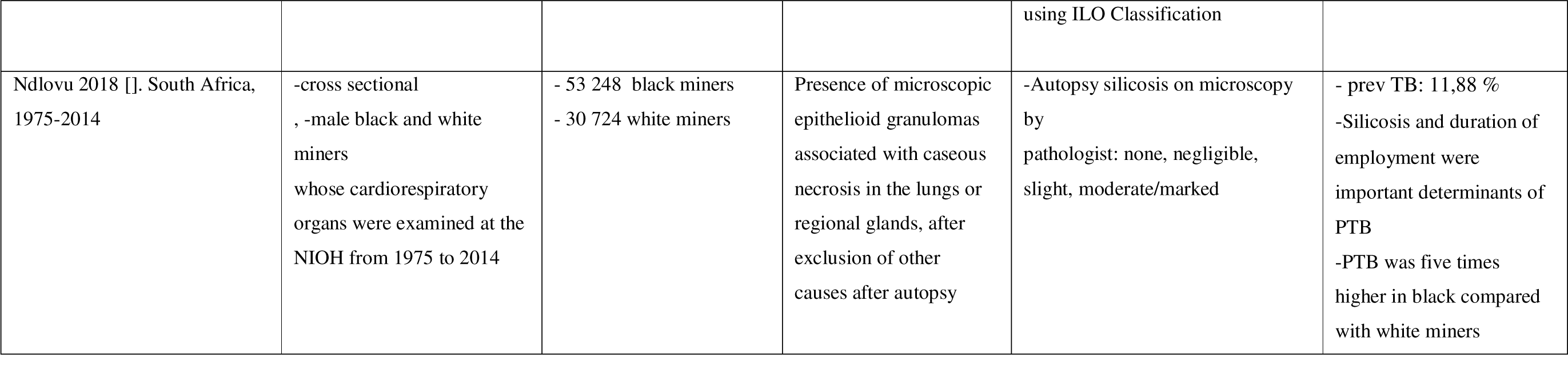
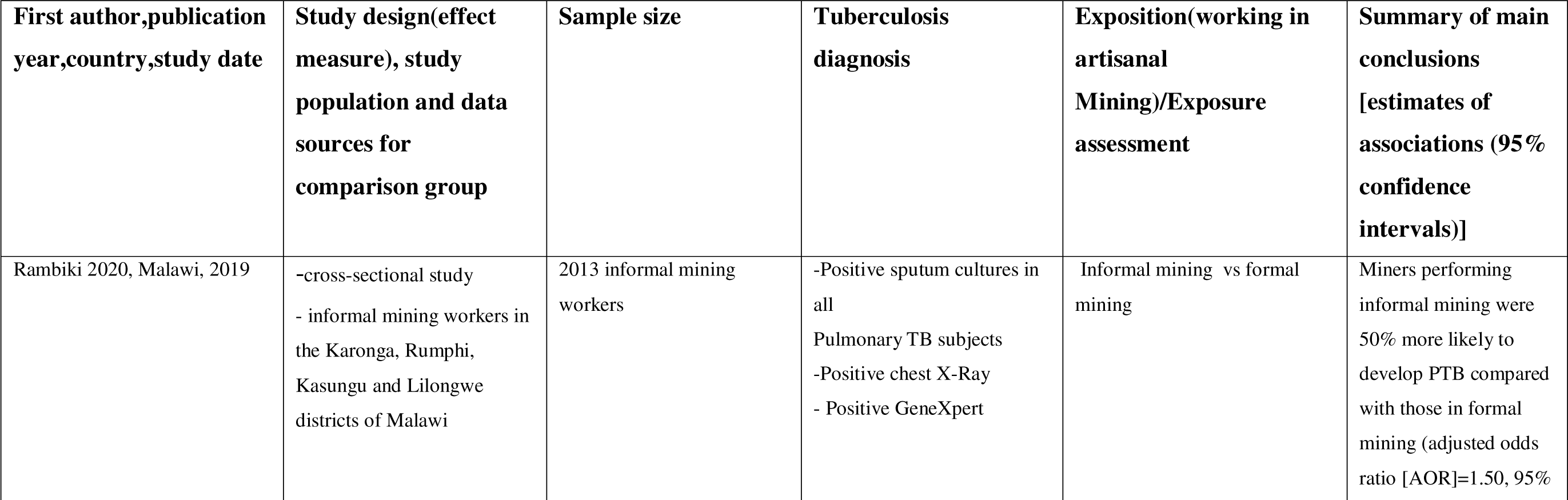
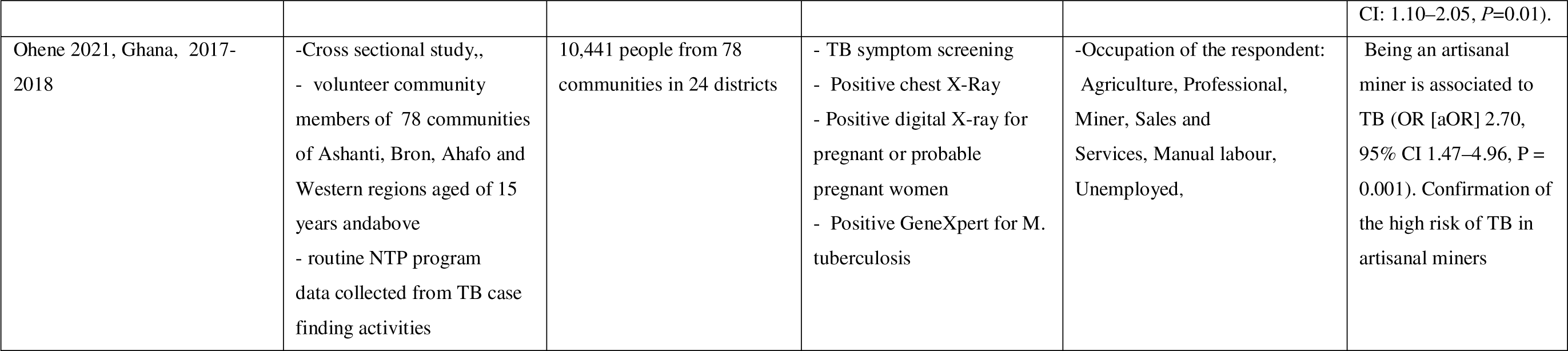
Artisanal mining and Tuberculosis: characteristics, sample size, and definition of exposure and outcome.

### 3.3 Overall situation of ASM miners, Incidence of TB and ASM-TB studies

Figure 2a depicts a map of SSA countries based on the number of ASM miners and TB incidence (new and relapse cases per 100,000 population per year). Out of the 22 SSA countries with over 100,000 ASM miners, five countries (DRC, Central African Republic, Angola, Mozambique, and Liberia) are among the thirteen highly to severely TB endemic SSA countries. However, none of these five countries have conducted a study on the association between ASM dust exposure and TB incidence, as demonstrated in Figure 2b which shows a map of SSA countries by number of ASM miners and ASM-TB studies.

**Fig.2a:**
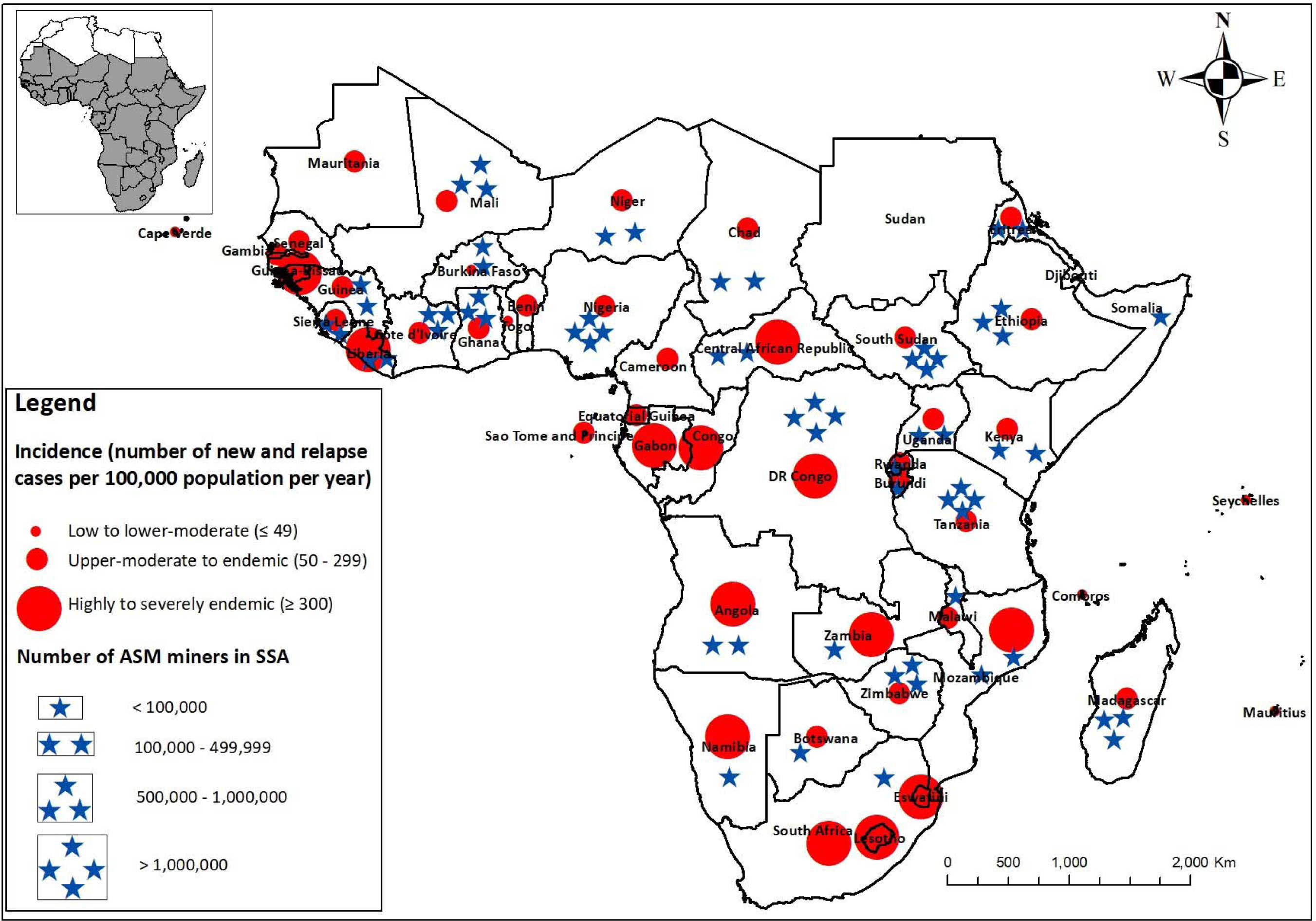
Map of SSA Countries by number of ASM miners and TB incidence (new and relapse cases per 100,000 population per year)

**Fig.2b:**
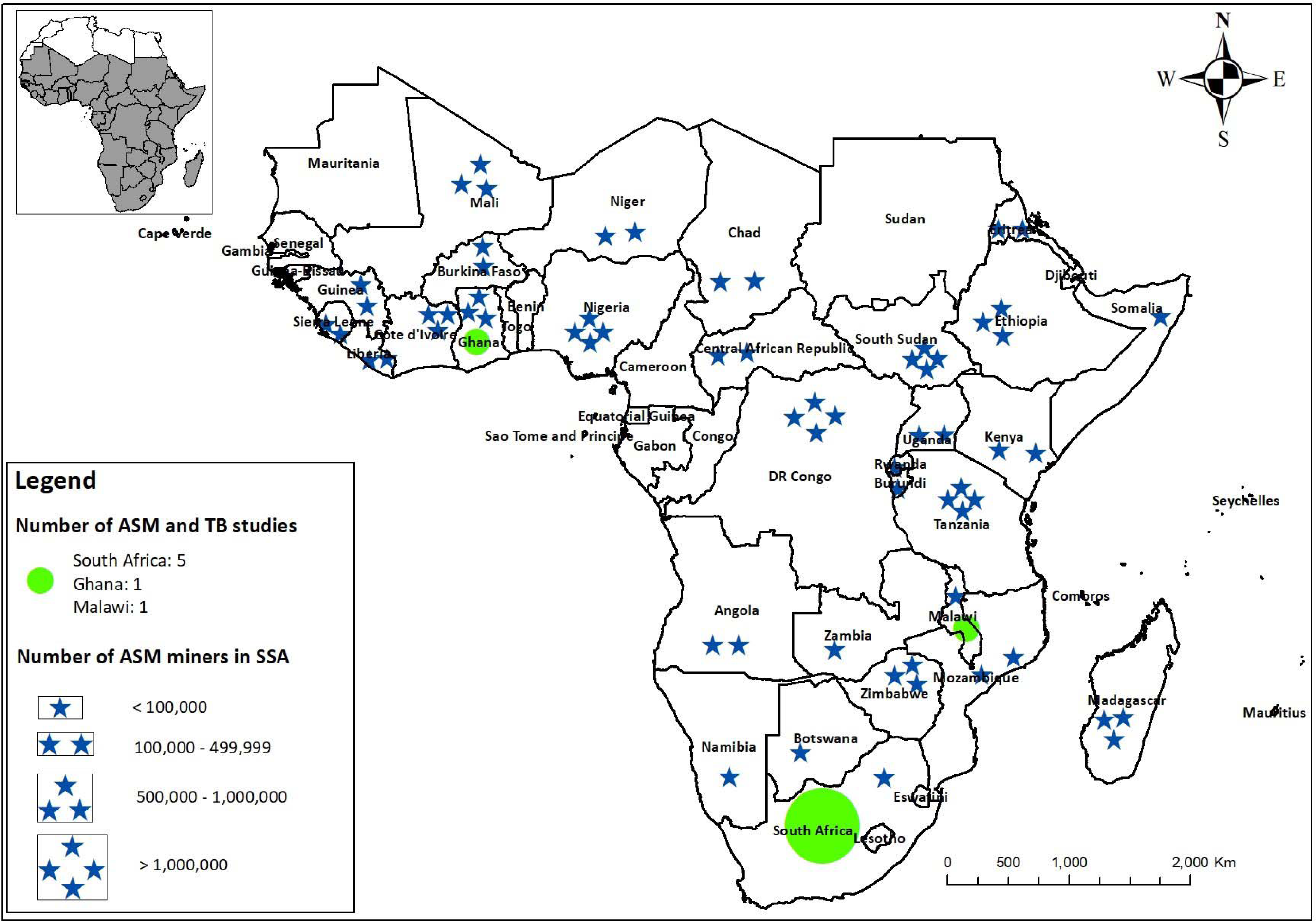
Map of SSA Countries by number of ASM miners and Tuberculosis-ASM studies.

**Fig. 3:**
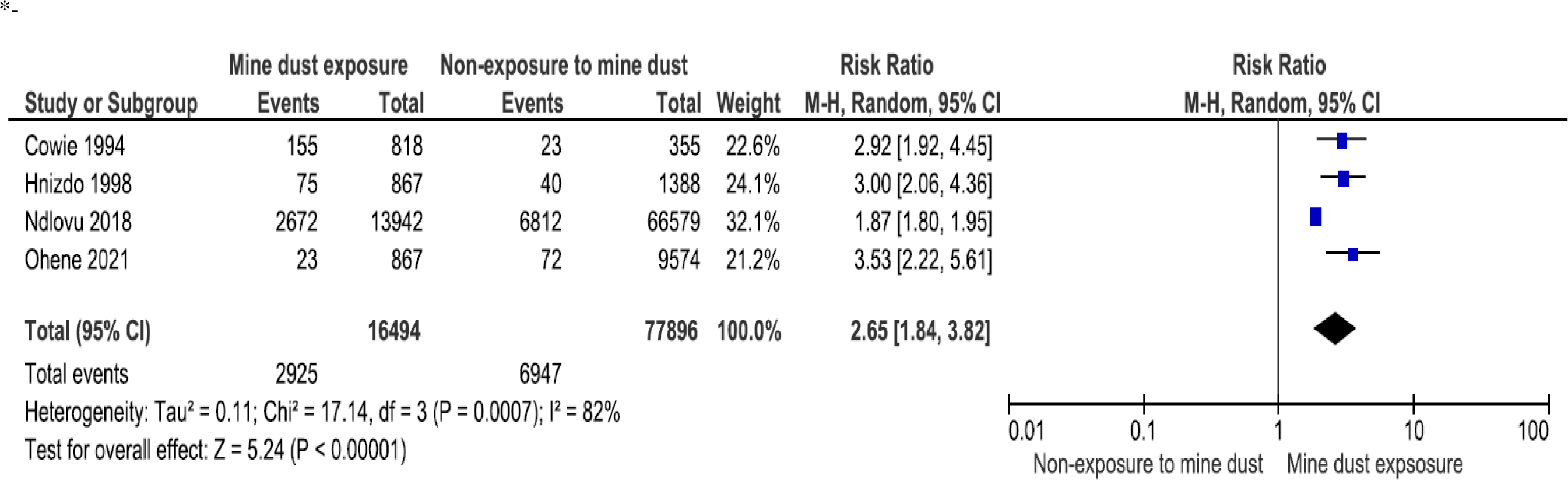
Meta-analysis results for mine dust exposure and incidence of tuberculosis.

### 3.4 Risk of bias assessment

Supplementary Tables 4a, 4b, and 4c (S3Tab4a,S3Tab4b and S3Tab4c)provided a summary of the risk of bias for the seven studies included in our analysis. With the exception of the comparability of study groups regarding additional factors beyond age (interpreted here as controlling for relevant additional factors), all studies were rated as having a low risk of bias. We also conducted tests to evaluate publication bias and found that it was less likely for TB prevalence among artisanal miners. Egger’s regression analysis showed no evidence of publication bias (z = 1.87, P = 0.061). However, Begg’s rank correlation analysis yielded a z-test of 0.38 (p = 0.707) (S2Fig2).

### 3.5 Exposure measurement and TB diagnosis

Four out of the seven studies [5, 19–21] used X-ray diffraction to measure silica dust exposure by Quartz fractions. They calculated the cumulative respirable dust and quartz exposures for each miner and used radiographs to diagnose Silicosis using the International Labor Organization (ILO). All included studies utilized standard methods for TB diagnosis except for one [21] which relied on clinical symptoms and Positive Chest X-Ray (CXR) for TB diagnosis. For instance, two out of the seven studies diagnosed TB based on positive sputum culture, positive CXR, and GeneXpert [23,24]. Other studies used either histopathology only [22] or CXR combined with positive sputum culture [5, 19]. Only one study combined CXR, positive sputum culture, and histopathology [20] (refer to Table 1).

### 3.6 Incidence of TB and silica dust exposure in artisanal/small scale mining

Seven studies were included in the analysis and all of them reported TB incidence in artisanal and small-scale mining (ASM). In South Africa, the incidence ranged from 5% to 35%. However, in Ghana and Malawi, the incidence was respectively 2.5% and 14%. A pooled analysis of data from these studies revealed that silica dust in ASM significantly increased the risk of TB incidence [RR 2.65 (95% CI: 1.84, 3.82)], with high heterogeneity among studies (P < 0.1, I2 = 82%) (Figure 4). The overall prevalence of TB among artisanal miners was estimated to be 15% (95% CI: 8.0, 23.0), with a pooled prevalence of 19% (95% CI: 11.0, 28.0) in high TB burden countries and 8% (95% CI: 3.0, 19.0) in upper-moderate TB burden countries. The test of group difference between countries with high and upper-moderate TB burdens was not statistically significant (P = 0.11) (Figure 8). Additionally, the meta-regression showed that a higher number of artisanal miners in Ghana was associated with a 1.64% reduction in TB incidence (P = 0.014) (S4Table 5).

**Fig. 4:**
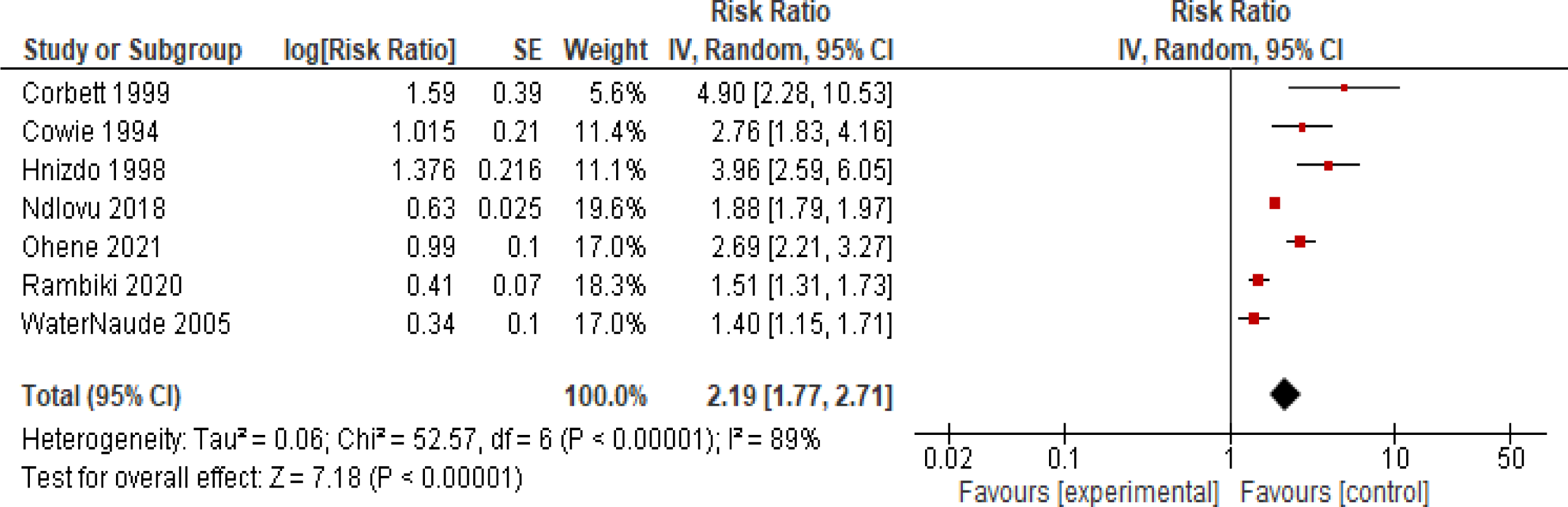
Estimate ratio (RR or OR) of tuberculosis in all the seven studies.

### 3.7 silica dust exposure in ASM and risk of TB

All studies included in the analysis consistently found that exposure to silica dust is the main risk factor for TB among artisanal miners. The cohort studies reported a relative risk (95% CI) ranging from 2.8 (1.9, 4.1) to 4.18 (2.75, 6.36), with a p value < 0.005. Case-control studies yielded an odds ratio of 4.90 (2.32, 10.58), while cross-sectional studies showed an odds ratio ranging from 1.50 (95% CI: 1.10, 2.05, P=0.01) to 35.2% (95%CI: 31.1, 39.3). A pooled analysis of the 7 studies confirmed that silica dust exposure was a significant risk factor for TB in ASM, with an estimate odds ratio of 2.19 (95% CI: 1.77, 2.71; I2 = 89%) (Figure 4). Furthermore, a subgroup analysis of the studies by design revealed a significant increased TB risk among those exposed to silica dust in ASM, with an estimate odds ratio of 2.24 (95%CI: 1.81, 2.78; I2 = 90%). Cohort studies showed a higher pooled odds ratio of 3.29 (95%CI: 2.31, 6.9, I2 = 90%), while cross-sectional studies had a pooled odds ratio of 1.80 (95%CI: 1.46, 2.22; I2 = 90%) (Figure 6).

### 3.8 Influence of the study year, the study design and the risk of TB in ASM

The combined findings indicate that the risk of TB in ASM is influenced by the year in which the study was conducted, with an overall summary estimate of 2.19 (95%CI: 1.77, 2.71) (Figure 5). However, subgroup analyses revealed a higher risk of TB among studies conducted before 2000 than those conducted after 2000, with summary relative risks (RRs) of 3.49 (95%CI: 2.57, 4.74) and 1.8 (95%CI: 1.46, 2.22), respectively. The level of heterogeneity among studies conducted before 2000 was minimal (P = 0.3, I2 = 15%), whereas among studies conducted after 2000, the heterogeneity was substantial (P < 0.1, I2 = 90%).

**Figure 5:**
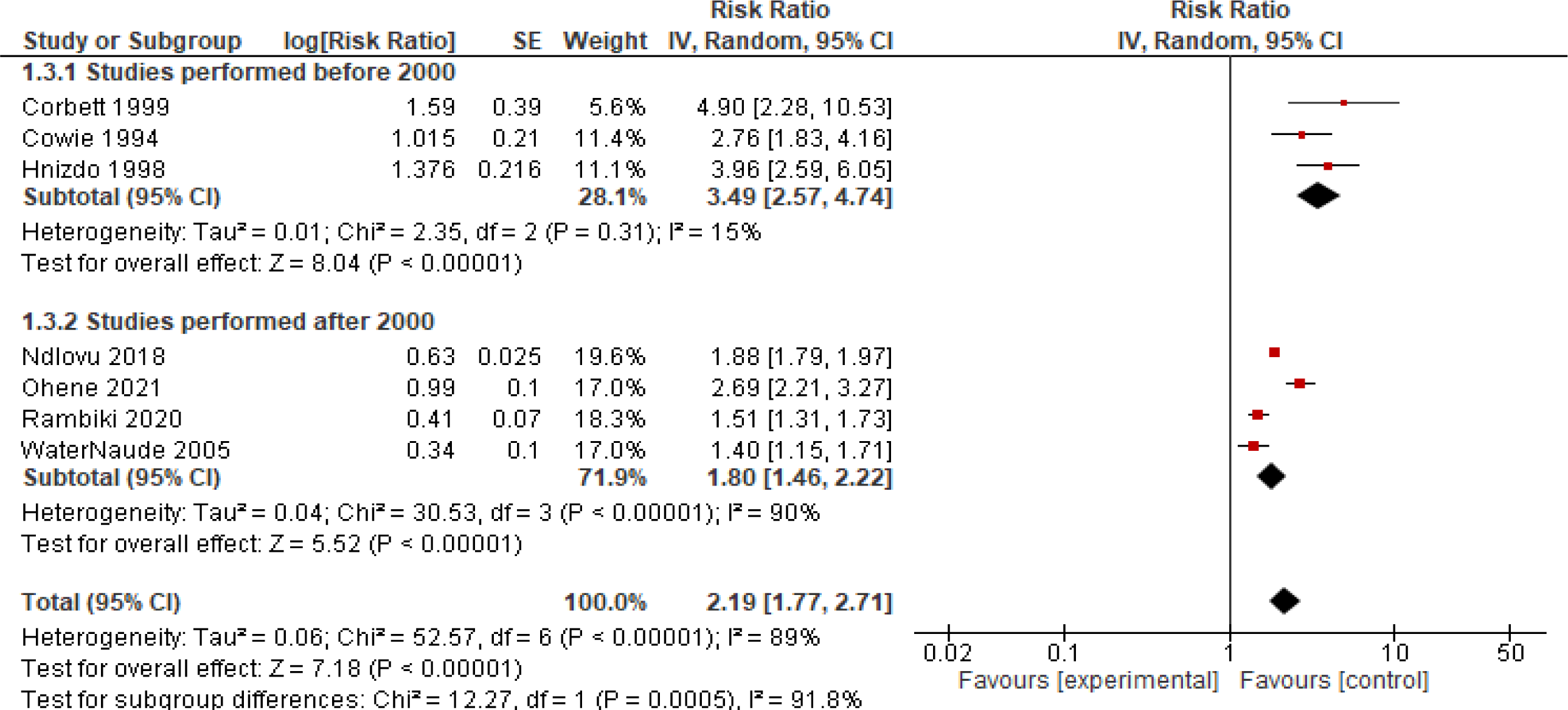
Estimate ratio of tuberculosis by study year.

**Figure 6:**
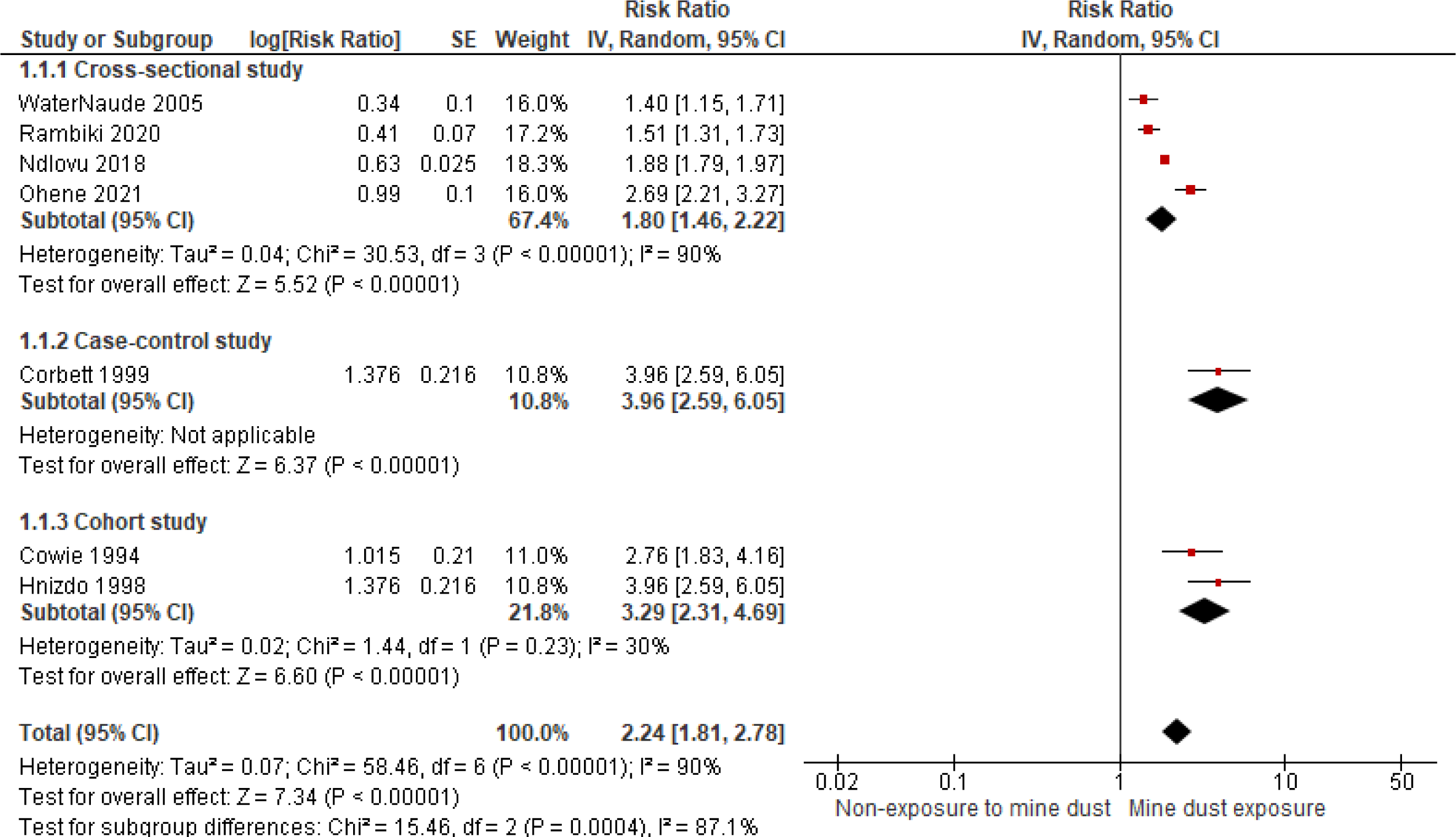
Estimate ratio of tuberculosis by study design.

### 3.9. Influence of high vs upper-moderate TB burden countries on TB disease among artisanal miners

The pooled prevalence of TB among artisanal miners was 19% [11%-28%] in high TB burden countries versus 8% [3%-19%] in upper-moderate TB burden countries. The overall prevalence of TB in artisanal workers was estimated to be 15% [8%-23%] (Figure 8). This is significant because the test of group difference was not statistically significant between countries with high and upper-moderate TB burdens (P = 0.11) (Figure 7).

**Fig.7:**
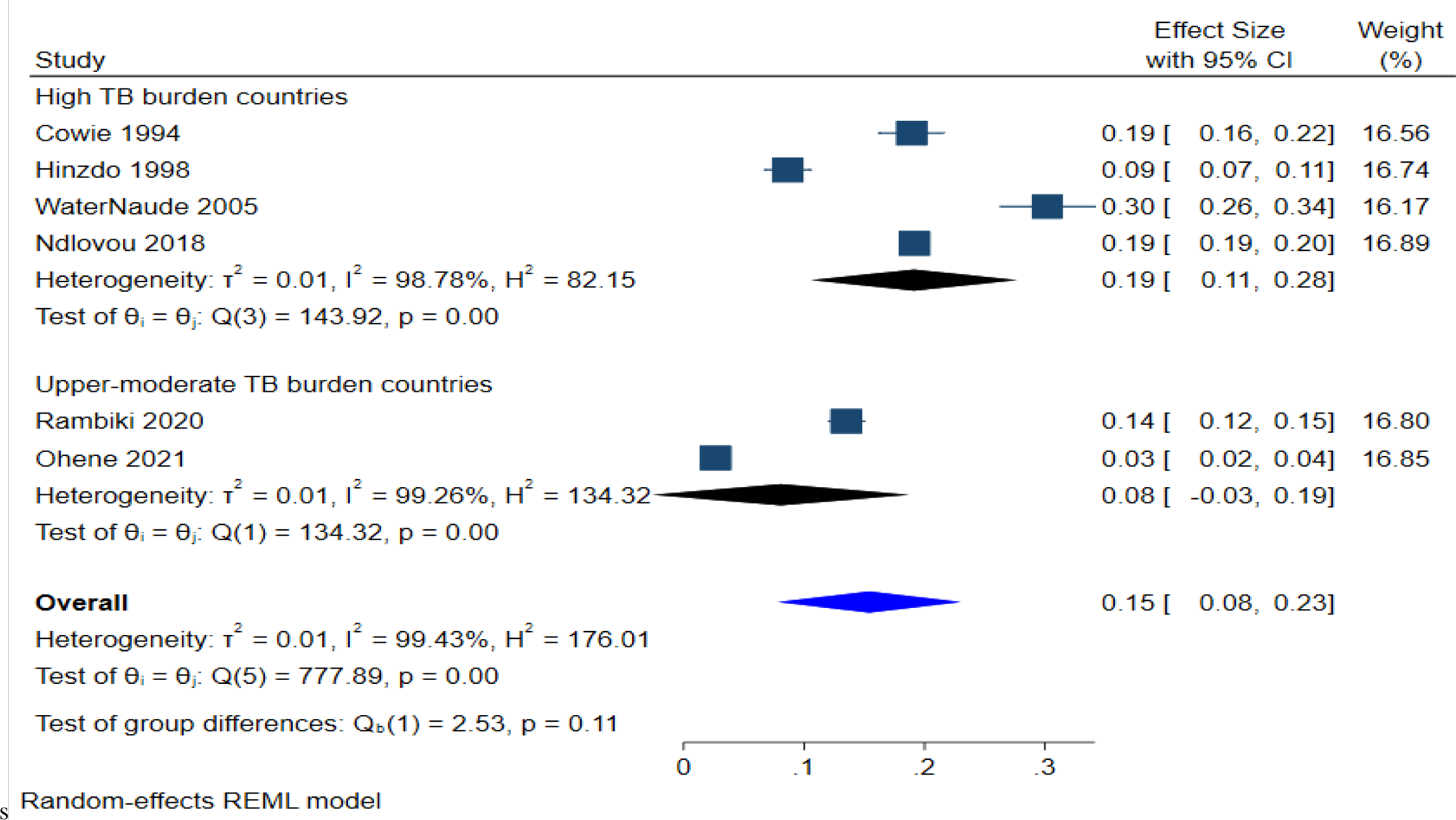
Influence of high vs upper-moderate TB burden countries on TB disease among artisanal miners.

## Discussion

### Summary of main evidence

This systematic review examines the association between silica dust exposure in ASM and the incidence of tuberculosis in Sub-Saharan Africa (SSA). Seven studies were identified and analyzed, covering only 3 countries out of the 46 in SSA, all with a high burden of tuberculosis. The review found a significant increase in TB incidence among those exposed to silica dust in ASM. The overall prevalence of TB in artisanal workers was estimated to be 15%, with a higher prevalence in countries with a high TB burden.

We also found that there was a significant increased risk of TB for exposed persons to silica dust in ASM no matter what the study design is (cohort or cross-sectional studies). Furthermore, this review revealed the influence of the study year on the risk of TB in ASM. In fact, this risk was higher in studies conducted before 2000 than in those conducted after 2000. This could be probably explained by the fact that most of the included studies were South African. Around the year 2000, voluntary industry milestones were established in South Africa to reduce 95% of all respirable crystalline silica dust measurements to below the occupational exposure limit (OEL) of 0.1 mg/m3 and to ensure no new cases of silicosis in novice miners entering the industry after 2008 [26]. These aim to reduce TB incidence to the national rate or below, and improve access to HIV counselling and testing assessment to antiretroviral therapy (ART), and also enhance health education and communication [27].

### Agreements and disagreements with other studies and reviews

The use of cumulative exposure measurement to silica dust and radiological silica exposure by ILO system agree with other findings elsewhere [32]. In California (USA), Checkoway et al. [35] has also the same methods to measure silica dust exposure. This method was also used by Taguchiet al. [36] to assess the relationship of mixed dust fibrosis and TB. With regards to the prevalence of TB, the high prevalence is consistent with findings in India by Chakraborty et al [37] who reported the TB prevalence rate in mining at 5.5%. The difference observed in different included studies may be explained by the diversity of TB diagnostic methods used in each study. For example, Waternaude et aL used clinical symptoms and Positive Chest X-Ray, Rambiki et al., and Ohene et al. used positive sputum culture, positive CXR and GeneXpert; Ndlovu et al. histopathology only; Cowie et al. and Corbett et al [5, 19] combined CXR and Positive sputum culture. Furthermore, the fact that some miners used to be underground and other on the surface may explain this difference as well.

This review found the existing association artisanal mining dust exposure and the risk of developing TB findings. This is consistent with other data from the literature. In fact, Stuckler et al. [31] has also found that mining production was associated with higher TB incidence rates in the population (adjusted b=0.093; 95%CI:0.067, 0.120); with an increased mining production of 1 SD corresponding to about 33% higher TB incidence or 760000 more incident cases), after adjustment for economic and population controls. This is also consistent with findings of Ehrlich et al. 2021, who investigated silico tuberculosis and dust exposure risk factors. He also concluded that occupational inhalation of silica dust increases the risk of pulmonary TB in co-exposed populations. In the same way, other studies, at the beginning of the 20^th^ Century, performed animal experiments on silica exposure and trabecle bacilli [38–40], and Mavrogordato. These studies concluded that silica dust exposition increases a great proliferation of bacilli and accelerates TB development.

Higher number of artisanal mines in Ghana was associated with 1.64% reduction of TB among artisanal miners. This can be explained by the improved policy of supervision of artisanal miners in Ghana [41]. Furthermore, the reduction in artisanal sites in favor of industrial exploitation would explain the reduction in TB cases compared to other countries in SSA. Recently, other findings in histological changes have reinforced this evidence. In his study, Pasula et al. [42] has demonstrated that silica pre-exposure for 30 days or more converts mycobacterial-resistant C57BL/6 mice into mice that are highly susceptible to both *M. avium* and *M. tuberculosis* infection. One of the included studies has reported the association of rifampicin and dust silica exposure in Ghana [24] and the resistance rate was 5.3%. These findings are in agreement with data from the National Surveillance data in Ghana [43] which reported drug resistance in artisanal miners. However, these findings are in disagreement with data provided by [44] Churchyard et al. who did not find association with mining activities and drug-resistance TB. Although there is the existence of some disagreements, that drug-resistance found in this review draws our attention to the fact that exposure to dust silica in mining is not only a factor associated with TB but could also impact on its treatment. It would then increase the burden of death.

This review has highlighted the scarcity of data in SSA on TB and artisanal or small-scale mines. In fact, studies on mining have been conducted in other SSA high, more specifically in middle burden of TB countries such as Tanzania [28, 29], DRC [12, 19], Nigeria [28] and in Ghana [30]. However, none of studies focused on TB as a health outcome among artisanal miners. Furthermore, a review [31] summarized literature on mining, a but did not include TB occurrence in artisanal miners. Our findings are however in the line with a review conducted by Ehrlich et al. [32] who addressed the risk factors of silicosis and TB in mining in the world. This review also mentioned scarcity of studies in high burden TB countries. In light of these findings, it has been noted that, to the best of our knowledge, no other studies have been found in countries such as the DRC, Ethiopia, and Burkina Faso, which are known to have the highest level of artisanal mining activities in the world, trailing only India, China, and Indonesia [33]. Furthermore, many other countries, including some of the previously mentioned are among the top 30 of countries in the world with a high TB burden [34]. This is the case of Angola (top one), Congo-Brazzaville, DRC, Ethiopia, Lesotho, Liberia, Mozambique, Namibia, Nigeria, Sierra Leone, Zambia and Zimbabwe. Although these listed countries have a high or intermediate level of artisanal or small-scale mining activities and are known to have a high or intermediate burden of TB, we know next to nothing about these big issues. Thus, studies assessing the risk factor of TB in artisanal mining areas should be conducted in countries such as DRC where we found numerous artisanal mining throughout the country and where no study for that concern has been conducted yet. Since this shortage of studies is established, data from these countries are needed in order to influence protective policies with different stakeholders.

#### Potential biases in the review process

To control potential biases in the review process, we used a structured and transparent approach. Inclusion criteria were set and grouped according to the PECO framework [14]. Structured searches of PUBMED, EMBASE and Scopus were undertaken using a combination of keywords. A structured tool for assessment of risk of bias was applied independently by two subject experts and agreed by consensus (NOS). We conducted publication bias tests to quantify publication bias. We found that publication bias was less likely for TB prevalence among artisanal miners when we ran statistical tests, and there was no evidence of publication bias in Egger’s regression analysis with z = 1.87 (P = 0.061). In contrast, Begg’s rank correlation analysis yielded a z-test of 0.38 (p = 0.707).

### Strengths and Limitations

Our study is the first systematic review conducted in Sub-Saharan Africa (SSA) that focuses exclusively on Artisanal and Small-scale Mining (ASM), which adds to the existing knowledge on this topic. However, despite our efforts, there are some limitations to our study that should be considered. Firstly, our review included observational studies, which are subject to selection bias. This means that the studies may not accurately represent the true prevalence of tuberculosis (TB) among artisanal miners in SSA, as they may not have included a representative sample of this population. Secondly, our review was limited by the number of studies available in the region. Due to a lack of studies, we were forced to exclude many countries from our analysis, which may have affected the generalizability of our findings. Additionally, some studies may have been missed due to language restrictions, which may have further limited the scope of our review. Despite these limitations, our review minimized publication bias, as we included studies from a variety of sources, including grey literature. Our findings are consistent with those conducted in other parts of the world, suggesting that the health risks associated with ASM are universal and require urgent attention. Finally, our study provides important evidence that can serve as a foundation for future health research on ASM and for advocacy to develop and implement health and environmental measures that can help to reduce the risks associated with ASM, and move towards a TB-free world in a mining environment.

## Conclusion

The prevalence of TB in artisanal mining activities is significant, posing a major risk not only to the miners but also to the wider community. This burden is expected to be twofold, impacting both the miners themselves and the communities in which they reside. Despite this, studies investigating this issue are scarce, particularly in regions of high and intermediate TB burden in SSA. Further, well-designed studies are needed to enhance our understanding of this issue. It is crucial that individual and governmental policies are implemented, including through national TB prevention and control programs, to improve working conditions and promote healthier societies.

### Box 1.

#### Evidence before this study

Exposure to silica data is known to be associated with high risk of developing tuberculosis. Most evidence comes from industrial mining activities. However, evidence in ASM including studies from SSA in lacking.

#### Added value of this study

This systematic review presents the available evidence on the association between artisanal mining and tuberculosis in SSA. As far as we are aware, this is the first systematic review on this topic in the region. The review confirms that artisanal mining is a significant risk factor for TB and remains a major public health challenge in SSA. The review also highlights that TB rates are high in artisanal mining and the emergence of drug-resistant TB is possible. However, the limited number of studies in this field reveals that there is a significant gap in knowledge on this topic, especially in many SSA countries such as the DRC where artisanal miners are likely to have some of the highest TB rates in Africa. Urgent research is required to address this issue and inform policies and interventions to improve the health and working conditions of artisanal miners in the region.

#### Implication of the study and recommendations

Based on the findings of this review, it can be concluded that artisanal mining activities have a significant impact on the burden of TB in SSA, leading to negative social and economic consequences. Therefore, urgent action is needed, ranging from personal protective measures to government policies aimed at creating healthy working environments. For instance, reducing poor working conditions, providing better living quarters, and limiting exposure to silica dust are essential measures that should be implemented [30]. Emphasis should be placed on countries with a high TB burden, particularly South Africa, where artisanal miners are at increased risk of TB infection. This highlights the need for strict TB preventive therapy (TPT) for this specific group and the establishment of clear policies on occupational medicine follow-up among artisanal miners. Additionally, it is essential to conduct studies to assess the risk factors of TB in artisanal mining areas in countries such as DR Congo, where numerous artisanal mining activities are present, and no studies have been conducted on this issue yet.

## Data Availability

All data produced in the present study are available upon reasonable request

## Acknowledgments

The authors are thankful to Josué MATABARO, Agricultural Engineer for the first design of the maps.

## Conflict of interest

The authors declare that they have no competing interests

## Funding

We received no funds for this review

## Human Participant Protection

No protocol approval was needed for this study.

## Availability of data and materials

The datasets used and/or analyzed during the current study are available from the corresponding author on reasonable request.

## Author’s contributions

Conceptualization: DGM, PK; Data curation: DGM, PMB, PK; Statistical analysis: RMK and JLT; Methodology: DGM, PK; Software DGM; map generation: FMZ; Supervision: PK; Writing—original draft, DGM; Writing—review & editing: DGM, PK, RMK, JLT, PMB, GAN,CM.

## Abbreviations

CI: confidence interval
OR: odds ratio
RR: relative risk, rate ratio or risk ratio
SIR: standardized incidence ratio
AM: Artisanal Mining
CXR: chest x-ray

## Supplementary files

**S1Table 2:**
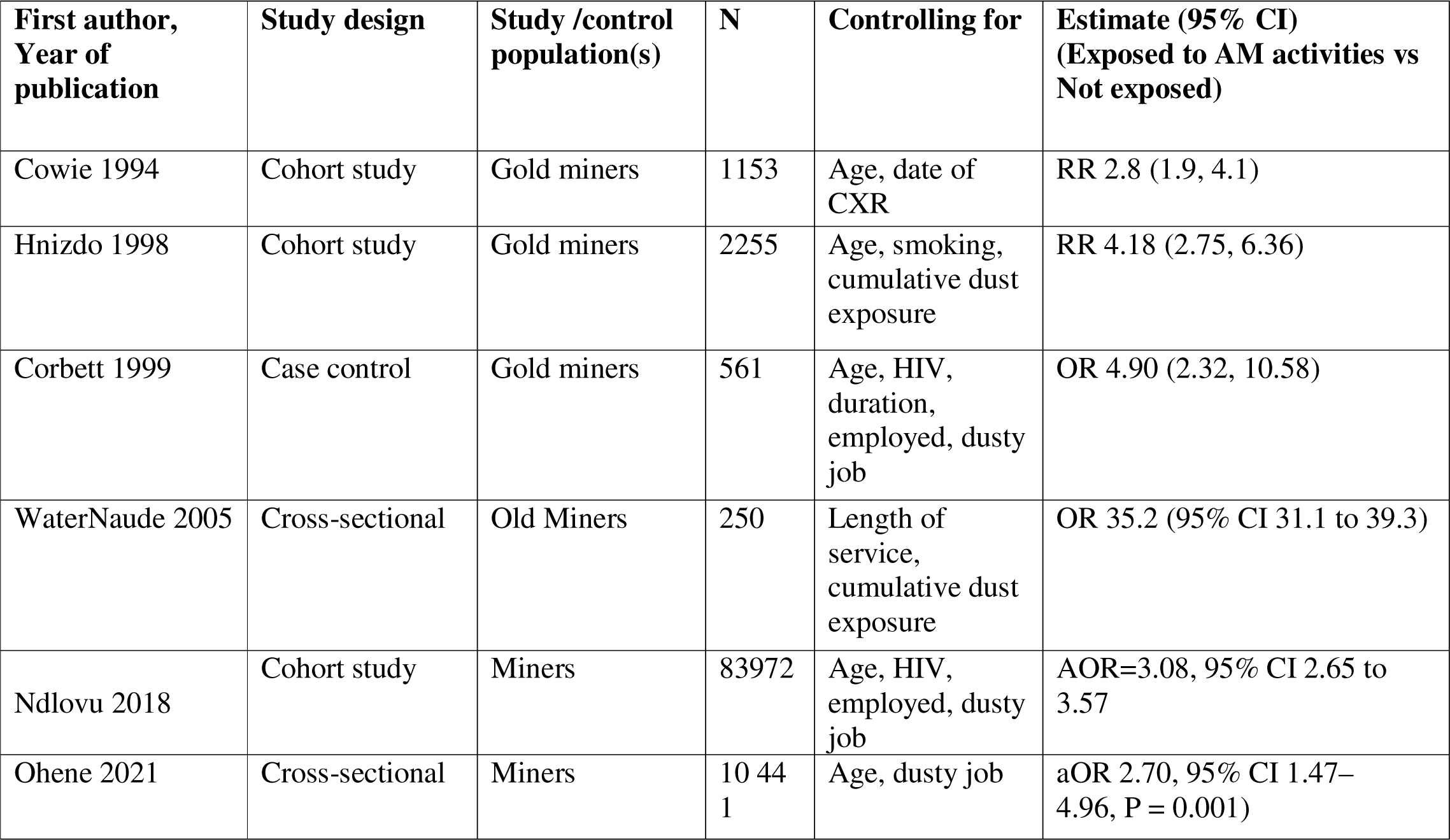
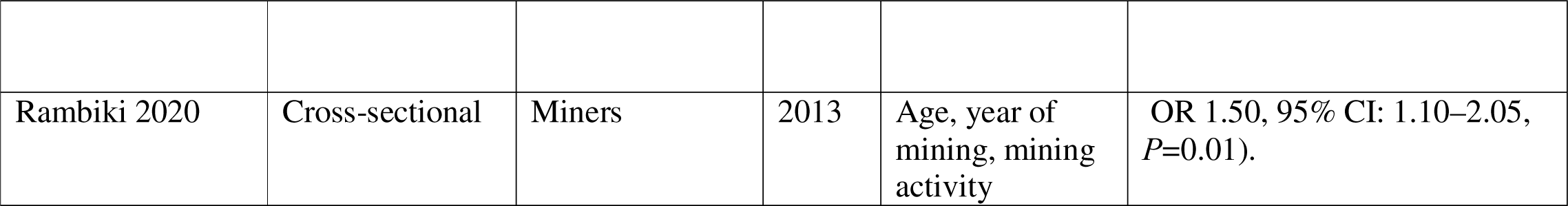
Prevalence of TB and Tuberculosis Risk or Odd by Artisanal Mining Exposure.

| **First author,**<br>**Year of**<br>**publication** | **Study design** | **Study /control**<br>**population(s)** | **N** | **Controlling for** | **Estimate (95% CI)**<br>**(Exposed to AM activities vs**<br>**Not exposed)** |
| --- | --- | --- | --- | --- | --- |
| Cowie 1994 | Cohort study | Gold miners | 1153 | Age, date of<br>CXR | RR 2.8 (1.9, 4.1) |
| Hnizdo 1998 | Cohort study | Gold miners | 2255 | Age, smoking,<br>cumulative dust<br>exposure | RR 4.18 (2.75, 6.36) |
| Corbett 1999 | Case control | Gold miners | 561 | Age, HIV,<br>duration,<br>employed, dusty<br>job | OR 4.90 (2.32, 10.58) |
| WaterNaude 2005 | Cross-sectional | Old Miners | 250 | Length of<br>service,<br>cumulative dust<br>exposure | OR 35.2 (95% CI 31.1 to 39.3) |
| Ndlovu 2018 | Cohort study | Miners | 83972 | Age, HIV,<br>employed, dusty<br>job | AOR=3.08, 95% CI 2.65 to<br>3.57 |
| Ohene 2021 | Cross-sectional | Miners | 10 44<br>1 | Age, dusty job | aOR 2.70, 95% CI 1.47–<br>4.96, P = 0.001) |
| Rambiki 2020 | Cross-sectional | Miners | 2013 | Age, year of mining, mining activity | OR 1.50, 95% CI: 1.10–2.05, *P*=0.01). |

**S2Table 3:** TB cases in exposed persons to silica dust versus non-exposed.

| Study ID | Exposed persons to the dust mines |  | Non-exposed persons to the dust mines |  | RR ou OR (95% CI) |
| --- | --- | --- | --- | --- | --- |
|  | Number of TB Cases | Total population | Number of TB Cases | Total population |  |
| Cowie 1994 | 155 | 818 | 23 | 335 | RR 2.8 (1.9, 4.1) |
| Hinzdo 1998 | 75 | 867 | 40 | 1388 | RR 4.18 (2.75, 6.36) |
| Corbett 1999 |  | 381 |  | 181 | OR 4.90 (2.32, 10.58) |
| WaterNaude 2005 | 157 | 520 |  |  | OR 35.2 (95% CI 31.1, 39.3) |
| Ndlovou 2018 | 2672 | 13942 | 6812 | 66579 | AOR 3.08, 95% CI 2.65 to 3.57 |
| Rambiki 2020 | 272 | 2013 |  |  | aOR 2.70, 95% CI 1.47– |
|  |  |  |  |  | 4.96) |
| Ohene 2021 | 23 | 867 | 72 | 9574 | OR 1.50, 95% CI: 1.10–2.05) |

**S3Table 4a.** Detailed Newcastle-Ottawa Scale of each included cohort studies for risk of bias assessment.

|  | Selection |  |  |  | Comparability |  | Outcome |  |  |  | Risk of bias |
| --- | --- | --- | --- | --- | --- | --- | --- | --- | --- | --- | --- |
| Study | Representativeness of exposed cohort | Selection of non-exposed cohort | Ascertainment of exposure | Demonstration that outcome of interest was present at start of study | Adjust for the most important risk factors | Adjust for other risk factors | Assessment of outcome | Follow-up length | Loss to follow-up length | Total quality score |  |
| **Cowie 1994** | 1 | 1 | 1 | 1 | 1 | ? | 1 | 1 | 1 | 9 | Low |
| **Hnizdo 1998** | 1 | 1 | 1 | 1 | 1 | ? | 1 | 1 | 1 | 9 | Low |

**S3Table 4b.** Detailed Newcastle-Ottawa Scale of each included case-control studies for risk of bias assessment.

| Study | Case definition adequate CC | Case representative CC | Selection of controls CC | Definition of controls | Case control comparability on age | Case control comparability on others factors | Ascertainment of exposure/blinding CC | Same method ascertainment CC | Non-responses, Equals? CC | Decision Risk bias |
| --- | --- | --- | --- | --- | --- | --- | --- | --- | --- | --- |
| Corbett 1999 | 1 | 1 | 1 | 1 | 1 | ? | 1 | 1 | 1 | Low risk |

**S3Table 4 c.**
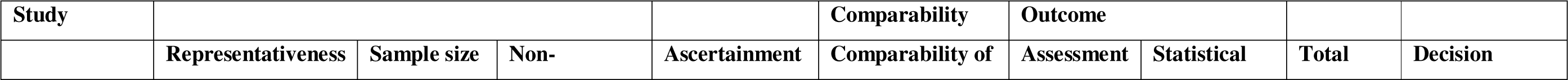
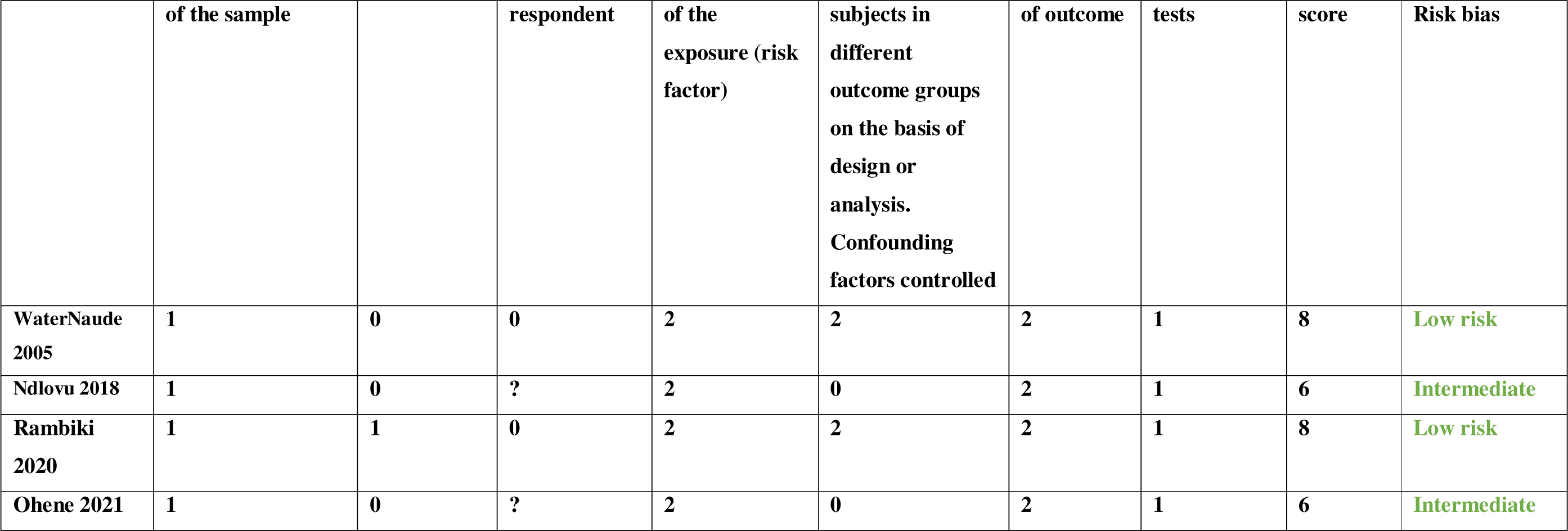
Detailed Newcastle-Ottawa Scale of each included cross-sectional studies for risk of bias assessment.

| Study |  |  |  |  | Comparability | Outcome |  |  |  |
| --- | --- | --- | --- | --- | --- | --- | --- | --- | --- |
|  | Representativeness | Sample size | Non- | Ascertainment | Comparability of | Assessment | Statistical | Total | Decision |

|  | of the sample |  | respondent | of the exposure (risk factor) | subjects in different outcome groups on the basis of design or analysis. Confounding factors controlled | of outcome | tests | score | Risk bias |
| --- | --- | --- | --- | --- | --- | --- | --- | --- | --- |
| WaterNaude 2005 | 1 | 0 | 0 | 2 | 2 | 2 | 1 | 8 | Low risk |
| Ndlovu 2018 | 1 | 0 | ? | 2 | 0 | 2 | 1 | 6 | Intermediate |
| Rambiki 2020 | 1 | 1 | 0 | 2 | 2 | 2 | 1 | 8 | Low risk |
| Ohene 2021 | 1 | 0 | ? | 2 | 0 | 2 | 1 | 6 | Intermediate |

**S4Table 5.**
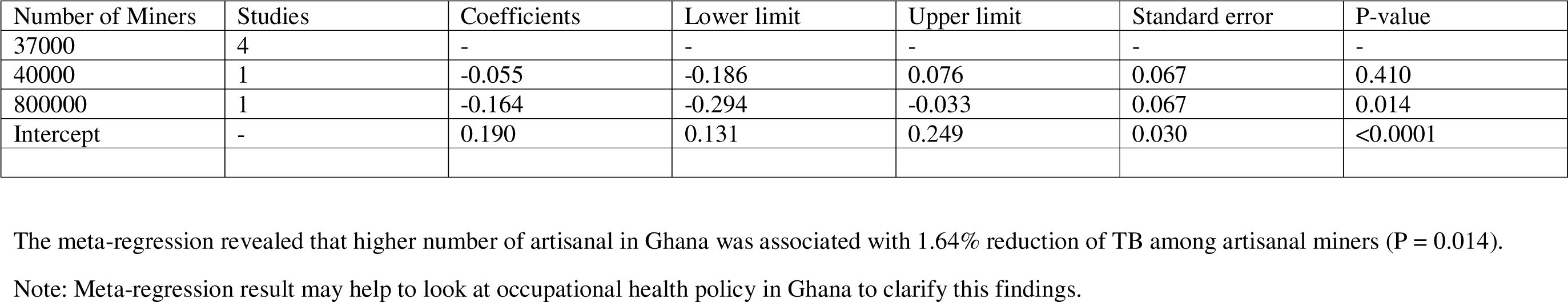
Meta-regression.

| Number of Miners | Studies | Coefficients | Lower limit | Upper limit | Standard error | P-value |
| --- | --- | --- | --- | --- | --- | --- |
| 37000 | 4 | - | - | - | - | - |
| 40000 | 1 | -0.055 | -0.186 | 0.076 | 0.067 | 0.410 |
| 800000 | 1 | -0.164 | -0.294 | -0.033 | 0.067 | 0.014 |
| Intercept | - | 0.190 | 0.131 | 0.249 | 0.030 | <0.0001 |

The meta-regression revealed that higher number of artisanal in Ghana was associated with 1.64% reduction of TB among artisanal miners (P = 0.014).
Note: Meta-regression result may help to look at occupational health policy in Ghana to clarify this findings.

### 1. Risk of bias assessment

**S3Table 4a**: Detailed Newcastle-Ottawa Scale of each included cohort studies for risk of bias assessment

The quality of included studies was assessed by the Newcastle Ottawa scale. A study can be awarded a maximum of one star for each numbered item within the Selection and Outcome categories and a maximum of two stars for Comparability.

#### Selection

1) Representativeness of exposed cohort: 1, study population truly or somewhat representative of a community/ population-based study; 0, study population was sampled from a special population, that is, population from a company, hospital patients, data from the health insurance company or health examination organization, nurses.
2) Selection of non-exposed cohort: 1, drawn from the same community as the exposed cohort.
3) Ascertainment of exposure: 1, Validation of pets use with secure record; 0, no specific pets use validation method.
4) *Demonstration that outcome was not present at start of study: 1, Comparability: 1) 1, whether a study adjusted for the most important factors deliberately; 1, whether a study adjusted for other important risk factors*.

#### Outcome

1) Assessment of outcome: 1, tuberculosis were confirmed by medical records or record linkage; 0, self-reported.
2) Was follow-up long enough for outcomes to occur: 1, duration of follow-up >= 5 year; 0, if duration of follow-up < 5 year.
3) Loss to follow-up rate: 1, complete follow-up or loss to follow up rate <=20 %; 0, follow-up rate < 80% or no description of those lost.

**S3Table 4b**: Detailed Newcastle-Ottawa Scale of each included case-control studies for risk of bias assessment

#### Selection

1) Is the case definition adequate?

a) Yes, with independent validation
b) Yes, e.g., record linkage or based on self-reports
c) No description
2) Representativeness of the cases

a) Consecutive or obviously representative series of cases
b) Potential for selection biases or not stated
3) Selection of Controls

a) Community controls
b) Hospital controls
c) No description
4) Definition of Controls

a) No history of disease (endpoint)
b) No description of source

#### Comparability

1) Comparability of cases and controls on the basis of the design or analysis

a) Study controls for age
b) Study controls for any additional factor

#### Exposure

1) Ascertainment of exposure

a) Secure record (e.g., pharmacy records)
b) structured interview where blind to case/control status
c) Interview not blinded to case/control status
d) Written self-report or medical record only
e) No description
2) Same method of ascertainment for cases and controls

a) Yes
b) No
3) Non-Response rate

a) Same rate for both groups
b) Non respondents described
c) Rate different and no designation

**S3Table 4 c.** Detailed Newcastle-Ottawa Scale of each included cross-sectional studies for risk of bias assessment

We used this adapted from the Newcastle-Ottawa Quality Assessment Scale for cohort studies to perform a quality assessment of cross-sectional studies

#### Selection

1) Representativeness of the sample:

a) Truly representative of the average in the target population. * (all subjects or random sampling)
b) Somewhat representative of the average in the target population. * (non-random sampling)
c) Selected group of users.
d) No description of the sampling strategy.
2) Sample size:

a) Justified and satisfactory. *
b) Not justified.
3) Non-respondents:

a) Comparability between respondents and non-respondents characteristics is established, and the response rate is satisfactory. *
b) The response rate is unsatisfactory, or the comparability between respondents and non-respondents is unsatisfactory.
c) No description of the response rate or the characteristics of the responders and the non-responders.
4) Ascertainment of the exposure (risk factor):

a) Validated measurement tool. **
b) Non-validated measurement tool, but the tool is available or described.*
c) No description of the measurement tool.

#### Comparability

1) The subjects in different outcome groups are comparable, based on the study design or analysis. Confounding factors are controlled.

a) The study controls for the most important factor (select one). *
b) The study control for any additional factor. *

#### Outcome

1) Assessment of the outcome:

a) Independent blind assessment. **
b) Record linkage. **
c) Self report. *
d) No description.
2) Statistical test:

a) The statistical test used to analyze the data is clearly described and appropriate, and the measurement of the association is presented, including confidence intervals and the probability level (p value). *
b) The statistical test is not appropriate, not described, or incomplete.

Comments: For quality and risk of bias assessment, we used the Newcastle-Ottawa Scale (NOS) (“Ottawa Hospital Research Institute,” n.d.), as recommended by the Cochrane Non-Randomized Studies Methods Working Group (“Cochrane Handbook for Systematic Reviews of Interventions,” n.d.). The NOS uses an eight-item rating system to evaluate the method of selecting participants, exposure/outcome assessment, and comparability among study groups. It has specific formats for cohort and case-control studies. We used the modified form for cross-sectional studies and case-control studies.

For cohort studies, a study could be assigned a maximum of one star for each numbered item within the selection and outcome categories and a maximum of two stars for comparability. The assessment was conducted as follows:

In the selection section, we assessed the representativeness of the exposed cohort, the selection of the non-exposed cohort, the ascertainment of exposure, the demonstration that outcome was not present at the start of study and assigned a score of 1 or 0 case by case.

1) Representativeness of the exposed cohort: 1, study population truly or somewhat representative of a community/ population-based study; 0, study population was sampled from a special population, that is, population from a company, hospital patients, data from the health insurance company or health examination organization, nurses.
2) Selection of the non-exposed cohort: 1, drawn from the same community as the exposed cohort.
3) Ascertainment of exposure: 1, Validation of pets use with secure record; 0, no specific pets use validation method.
4) Demonstration that the outcome was not present at start of study: 1, Comparability: 1) 1, whether a study adjusted for the most important factors deliberately; 1, whether a study adjusted for other important risk factors.

In the outcome section, we assessed whether TB was confirmed by one of the recommended methods or self-reported and assigned a score of 1 or 0 accordingly. Then we assessed the follow-up length for the outcome to occur and assigned a score of 1 if the duration of follow-up >= 5 year or 0 if the duration of follow-up < 5 year. Finally, we assessed the loss to follow-up rate and assigned 1 for complete follow-up or loss to follow up rate <=20 % and 0 for follow-up rate < 80% or no description of those lost.

For cross-sectional studies, we used an adapted format from the Newcastle-Ottawa Quality Assessment Scale for cohort studies to perform a quality assessment of cross-sectional studies (see additional files table 4c). In the Selection section (Maximum 5), we assessed the representativeness of the sample, the sample size, non-respondents, and ascertainment of the exposure (risk factor). In this point, we awarded for the sample representativeness 1 if the sample was truly representative of the average in the target population (all subjects or random sampling) or if it was somewhat representative of the average in the target population (non-random sampling) and 0 if the sample was from selected group of users or if there was no description of the sampling strategy. For the sample size, we scored 1 if it was justified and satisfactory and 0 if not justified. About non-respondents, we assigned 1 if the comparability between respondents and non-respondents characteristics is established, and the response rate is satisfactory and 0 if the response rate is unsatisfactory, or the comparability between respondents and non-respondents is unsatisfactory and if there was no description of the response rate or the characteristics of the responders and the non-responders.

For the ascertainment of the exposure (risk factor), we scored 2 if the study used a validated measurement tool, 1 if it used a non-validated measurement tool, but the tool was available or described, and 0 if there was no description of the measurement tool.

In the comparability section (maximum 2 scoring), we assessed whether the subjects in different outcome groups are comparable, based on the study design or analysis. If confounding factors are controlled or not. Then, we awarded 1 if there were study controls for the most important factor (select one) or if there was a study control for any additional factor and 0 if not.

In the outcome section (maximum 3), we awarded a score of 2 independent blind assessment of the outcome and the record linkage; a score of 1 for self-report; and 0 if no description was provided. In the same section, we assessed statistical test and assigned 1 if the statistical test used to analyze the data was clearly described and appropriate, and the measurement of the association was presented, including confidence intervals and the probability level (p value). In case that the statistical test was not appropriate, not described, or incomplete; we assigned a score of 0.

**S5Table 6.**
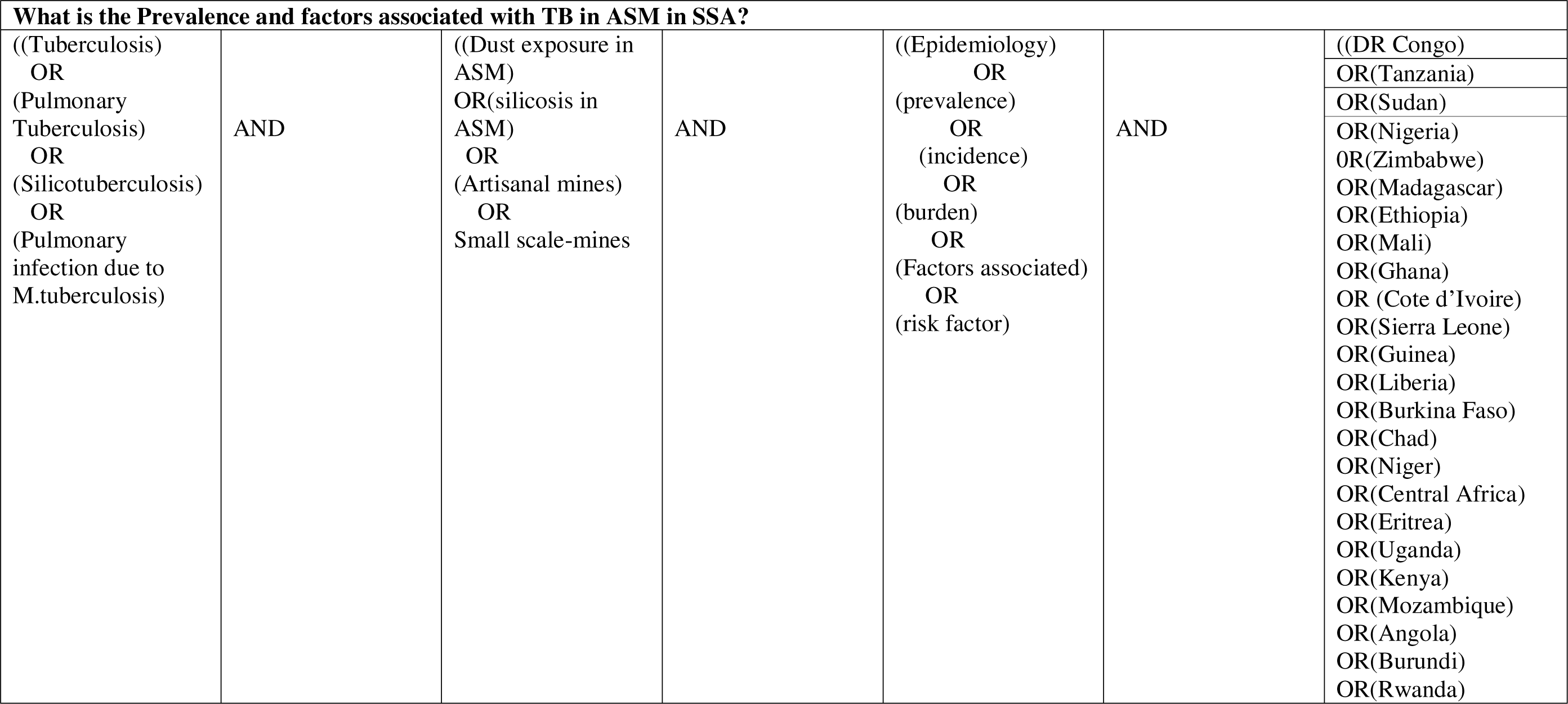
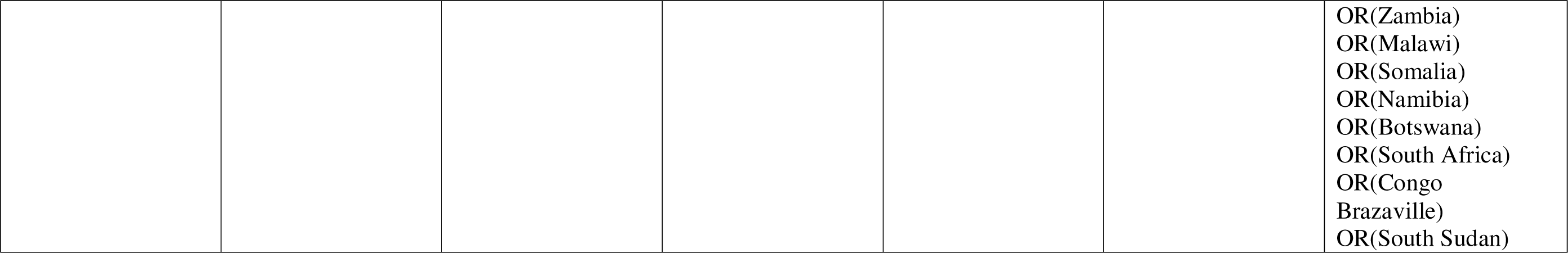
Surch strategy according to PICO Question.

**S1Figure 1.**
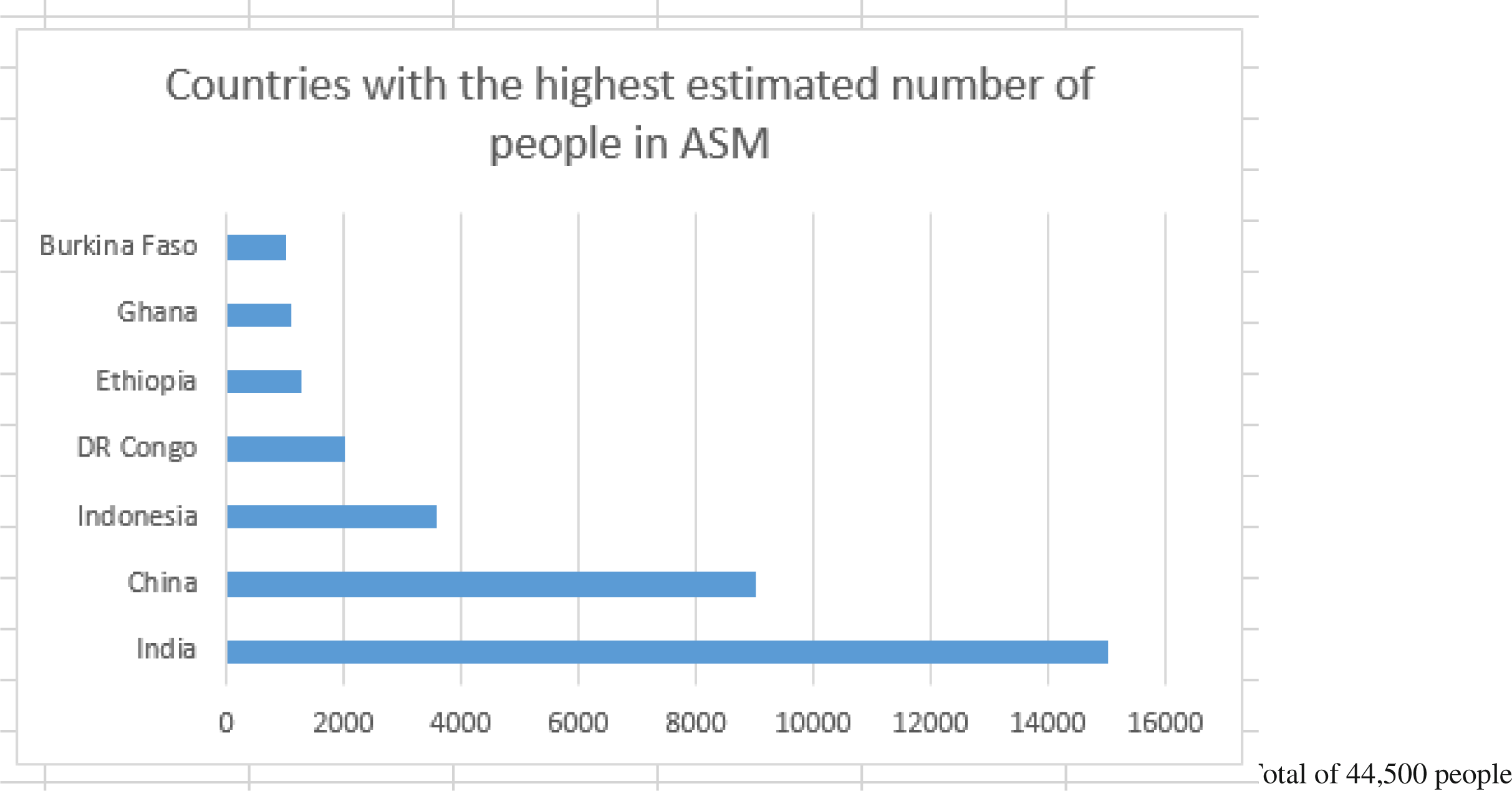
Countries with the highest estimated number of people working in artisanal and small scale mining (https://www.statista.com)

**S2Fig 2:**
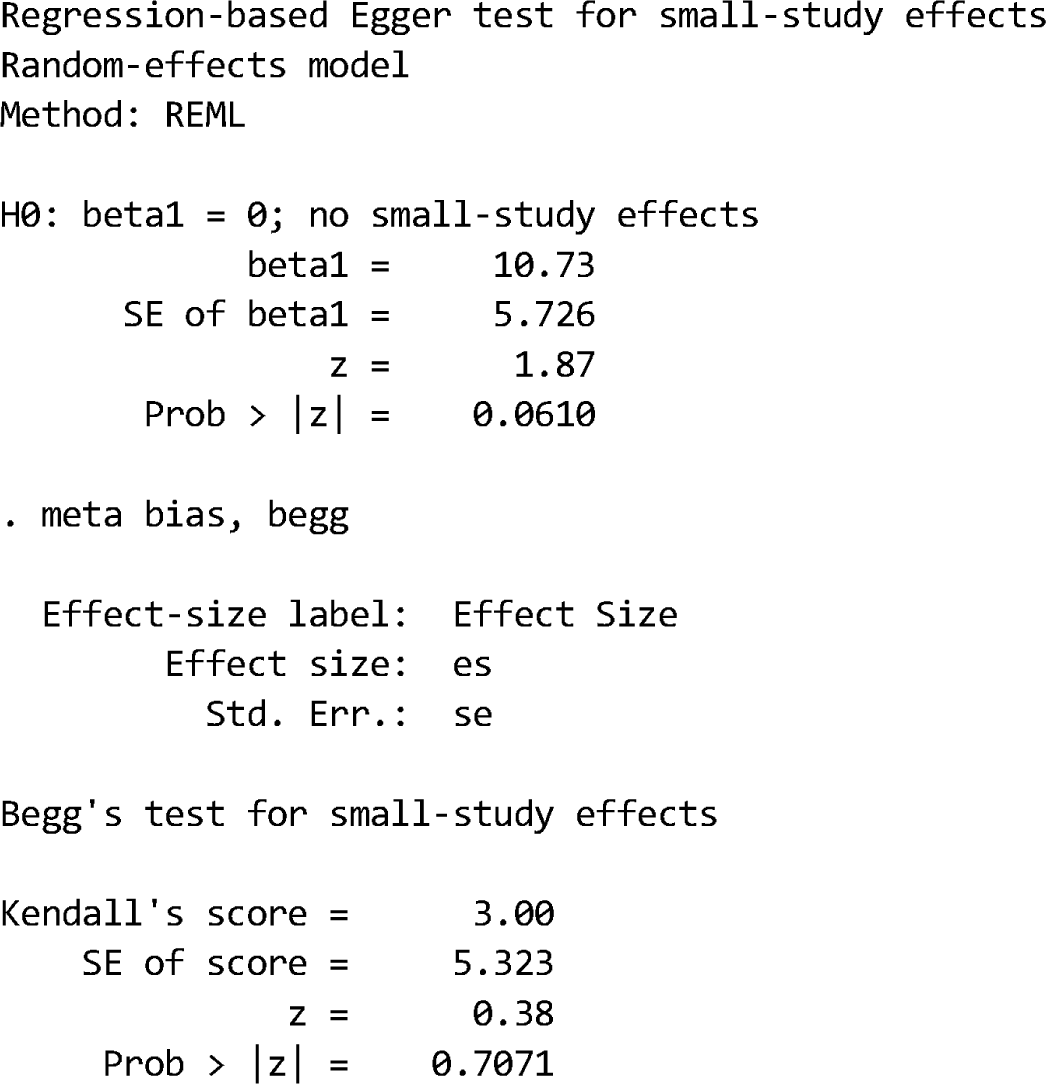
Publication bias.

## Notes

### Competing Interest Statement

The authors have declared no competing interest.

### Funding Statement

This study did not receive any funding

